# Academic dependency: the influence of the prevailing international biomedical research agenda on Argentina’s CONICET

**DOI:** 10.1101/2022.05.12.22275000

**Authors:** M. García Carrillo, F. Testoni, M. A. Gagnon, C. Rikap, M. Blaustein

**Affiliations:** Instituto de Biociencias, Biotecnología y Biología Traslacional (iB3), Departamento de Fisiología, Biología Molecular y Celular (DFBMC), Facultad de Ciencias Exactas y Naturales (FCEyN), Universidad de Buenos Aires (UBA), Buenos Aires, Argentina; Instituto de Lingüística, Facultad de Filosofía y Letras (FFyL), Universidad de Buenos Aires (UBA), Buenos Aires, Argentina; School of Public Policy and Administration, Carleton University, Ottawa, Canada; CITYPERC, City, University of London, London, United Kingdom; Consejo Nacional de Investigaciones Científicas y Técnicas (CONICET), Argentina

**Author notes:** Corresponding authors (CR), (MB). These authors contributed equally to this work (M. García Carrillo, F. Testoni). These authors also contributed equally to this work (C. Rikap, M. Blaustein).

## Abstract

**Background:** Previous research within the field of health and biomedical sciences (HBMS) reported that its prevailing research agenda is determined by leading academic institutions and big pharma companies, prioritizing the exploration of novel pharmacological interventions over research on the socio-environmental determinants of disease. Unlike previous studies, which have relied primarily on qualitative analyses, the aim of this investigation is to quantitatively explore if that prevailing international research agenda influences research in semi-peripheral countries and to which extent.

**Methods:** We used the Web of Science database and the CorText platform to proxy the HBMS research agenda of a prestigious research institution from Latin America: Argentina’s National Research Council (CONICET). We conducted a bibliometric and lexical analysis of 16,309 HBMS scientific articles whereby CONICET was among the authors’ affiliations. The content of CONICET’s agenda was depicted through co-occurrence network maps of the most prevalent multi-terms found in titles, keywords, and abstracts. We compared our findings with previous reports on the international HBMS research agenda.

**Results:** In line with the results previously reported for the prevailing international agenda, we found that multi-terms linked to molecular biology and cancer research hegemonize CONICET’s HBMS research agenda, whereas multi-terms connecting HBMS research with socio-environmental cues are marginal. However, we also found differences with the international agenda: CONICET’s HBMS agenda shows a marginal presence of multi-terms linked to translational medicine, while multi-terms associated with categories such as pathogens, plant research, agrobiotechnology, and food industry are more represented than in the prevailing agenda.

**Conclusions:** In line with the academic dependency theory, CONICET’s HBMS research agenda shares topics, priorities, and methodologies with the prevailing HBMS international research agenda. However, CONICET’s HBMS research agenda is internally heterogeneous, appearing to be mostly driven by a combination of elements that not only reflect academic dependency but also economic dependency.

## Introduction

The field of health and biomedical sciences (HBMS) constitutes a model case of blurred boundaries between academic and commercial research [1], and thus represents an ideal system to investigate the influence of corporate interest on research agendas. It has been shown that collaborations between industry and public research institutions generate a skewing problem (or dilemma), by creating a bias in academic agendas towards private needs [2,3]. It is well documented that conflicts of interest in medical research can influence research results [4–6]. Corporate interests can also drive research agendas away from those questions that are most relevant to public health [7].

It was also observed that large pharma companies typically sponsor and establish agreements with research institutions from core countries [8]. Globally recognized institutions from non-core countries, defined as semi-peripheral institutions, seldom call their direct attention; in particular, they are not among big pharmaceuticals’ frequent partners [8]. Furthermore, the academic literature agrees that there is a lack of demand for technology transfer in non-core countries. Latin America is characterized by a lack of new innovative firms [9] and most local corporations do not invest in Research and Development (R&D), being mostly demanders of technical assistance [10–12]. Therefore, we may (misleadingly) conclude that the skewing problem in research institutions from this region is absent. Since local companies do not tend to demand new research, direct contacts between local research institutions and global leaders are rare and, if they take place, they tend to be concentrated in a small group of internationalized research teams [13], it could be concluded that the bias of the overall local research agenda towards private needs will be negligible.

However, direct contacts between actors are one, but not the only way in which large private firms can influence public agendas. Indeed, beyond the acknowledgement of a direct influence of corporate interests in setting research agendas when corporations fund academic investigations or publish papers, Testoni et al. (2021) have recently shown that the influence of large pharma companies drives HBMS’ prevailing research agenda [14]. By performing a bibliometric and lexical analysis of more than 95,000 scientific articles published between 1999 and 2018 in the highest impact factor journals within HBMS, the authors found that the HBMS prevailing research agenda results from the intertwining of leading academic institutions and large private firms’ agendas. The resulting HBMS prevailing research agenda at the international level, as shown in that research, is more inclined towards molecular biology approaches and methodologies and cancer related research. Furthermore, it prioritizes investigations on pharmacological intervention over studies on the socio-environmental factors influencing disease onset or progression [14].

An open question in relation to these findings is whether this HBMS prevailing research agenda permeates into the priorities of institutions that do not actively participate in shaping it. Researchers from institutions that do not participate in defining the international research agenda may also emulate it, even if they share no direct link with the institutions that set it. The need to publish in internationally recognized journals, to remain or move forward in academic careers, and the perception of certain topics and methods as frontier science could be among the reasons why these other institutions adopt the prevailing international research agenda in HBMS. If this were the case, the reach of big pharma companies’ influence in setting research agendas would be even larger than what Testoni et al. (2021) found [14].

This question is even more pressing when considering institutions that might have other social and/or economic priorities. In the case of public research organizations from non-core countries, the international research agenda may compete (and eventually replace), complement, or co-evolve with an agenda focused on contributing to local priorities. In low- and middle-income countries, the latter could include a focus on promoting a different paradigm in science and technology that addresses urgent social and environmental issues [15–17].

The academic literature has coined the term “academic dependency” to explain the influence of international prevailing research agendas -mainly driven by core countries’ elite research institutions-on local research agendas, priorities, and the overall research of non-core countries’ academic institutions [15,18,19]. In its simplest form, the academic dependency approach suggests that both low and middle-income countries’ scientific research adopt that prevailing agenda [20], and that researchers from those countries occupy subordinate positions in science’s global division of labor [18]. Other authors, however, provide a more nuanced understanding of the interplay between international and local research agendas [15]. They argue that the concept of academic dependency is “attached to a simplifying perspective” of the relation between international and local scientific agendas, since it overlooks “the complexity of asymmetries that are located within the structure of a field crossed by international, regional and local circuits of recognition” [15]. In this same line, Beigel argues that research fields in the peripheries are internally heterogeneous [16]. Her work highlights the limitations of considering non-core countries as passive importers of knowledge. From that standpoint, the author concludes, “academic heteronomy and autonomy coexist in specific historical situations, which should be examined empirically” [16].

Overall, previous studies used qualitative methodologies, especially case studies, to assess academic dependency. These investigations do not account for the general impact of academic dependency on the definition of a local research agenda. To overcome this shortfall and to study the potential existence of an additional indirect skewing problem, we analyzed the research agenda of Argentina’s National Research Council (CONICET). The aim of this investigation was to quantitatively explore if the prevailing international research agenda, set by big pharma and leading academic institutions, indirectly influences academic research in non-core countries and to which extent. Given that CONICET is a case of an institution detached from the HBMS international research agenda but at the same time it is a recognized and prestigious institution in the field, it represents a good case for tackling this question.

We found that the CONICET’s HBMS research agenda is closely similar to the prevailing HBMS prevailing research agenda both by privileging certain diseases and favoring specific research approaches and methodologies. Nevertheless, our results also show that CONICET’s HBMS research agenda is internally heterogeneous, providing evidence of the co-habitation of academic heteronomy and a certain degree of autonomy. The presence of some distinctive research topics found in CONICET’s research agenda, which point to the presence of at least some aspects of academic autonomy vis-à-vis the prevailing HBMS research agenda, could be the result of Argentina’s asymmetries with core countries. Among them, the specific content of the CONICET’s HBMS research agenda could be explained by its role in the international division of labor (as a supplier of mostly primary goods) and specific health issues affecting Argentina’s population. In this sense, we conclude that the research agenda of a semi-peripheral institution like the CONICET seems to be the result of a combination of different factors that mostly reflects academic as well as economic dependency.

## Materials and Methods

In order to reconstruct CONICET’s research agenda in HBMS, we adopted the same methodology applied in Testoni et al. (2021) for building proxies of the content of HBMS prevailing research agenda [14]. Briefly, we extracted a corpus of HBMS scientific publications from the Web of Science (WoS) that included at least one CONICET author. Since the WoS classifies journals in terms of scientific categories, we manually picked out all the specific categories corresponding to HBMS. Next, we acquired the full list of journals that belonged to any of those specific categories and we retrieved all their available publications (including original research articles, reviews, perspectives, editorials, among others) that had been authored by the CONICET between 1999 and 2018. Overall, we retrieved 16,309 papers. To assess the temporal evolution of CONICET’s research agenda in line with the periodization proposed by Testoni et al. (2001), we divided the corpus of publications into two regular sub-periods: 1999-2008 and 2009-2018. CONICET’s publications grew 161% (from 4,523 to 11,789) in the second period under consideration [14].

The data were processed using the CorText platform [21], which allowed us to build co-occurrence maps by using specific algorithms that associate entities according to their frequency of co-occurrence within a chosen corpus of texts [22]. In our case, the corpus consisted of a CONICET scientific publications’ set, and we analyzed the prevailing content of the research included in it, proxying CONICET’s HBMS research agenda. We followed Tancoigne et al. [21] in their procedure to draw the maps, including the filtering of the corpus.

In this work, and regarding the definition of *agenda*, we appealed to the concept developed by Discourse Analysis that conceives it as a hierarchically ordered set of topics shared inside a particular community [23]. Following McCombs, we studied the content of the agendas rather than the efforts to dictate them (for instance, public policies regarding HBMS) [23]. In our definition, the agenda is also a reconstruction of shared representations and their values for each community [24]. For example, in the HBMS community, it includes the relevance given to a scientific issue, the validity of a method, or the objectivity of an approach. Therefore, network maps that interconnect contents of the agenda and show the centrality and the bridging position of a topic (to proxy its relevance) are an adequate methodology for studying agenda setting from a discourse analysis perspective.

We identified CONICET’s position in the ranking of organizations previously determined in Testoni et al. [14] to provide additional evidence of the fact that CONICET does not belong to the core of organizations that set the international HBMS research agenda. To reconstruct the CONICET’s HBMS research agenda, we performed a lexical analysis of the titles, keywords, and abstracts of the retrieved corpus of CONICET’s HBMS papers. We extracted the top 500 multi-terms of up to five words as a proxy of privileged research topics among HBMS and techniques associated with these topics. Monograms were excluded, and each list was refined following an in-depth cleaning process. This filtering was performed to avoid words not related to the HBMS field and whose frequency responds to either their grammatical function (such as “and” and “or”) or the level of grammaticalization within the scientific genre (“significance and impact”, “proposed method”, “detection limit”, etc.). The resulting list of 393 multi-terms was classified into general categories according to research topics (such as “Cancer/Tumor” or “Cardiovascular”). During the process of selecting the key multi-terms, we found that many of them referred to methods and procedures. Since they provide valuable data on the nature of the research under analysis, we decided to include them. For instance, a paper could contain terms such as “gel electrophoresis” or “western blot” in its title and/or abstract, which may be indicative of the fact that the topic is being studied from a molecular and cellular biology perspective and/or using tools associated with this field.

A network map was constructed for each period, plotting its corresponding most frequently connected multi-terms represented as nodes. We prioritized the top 100 multi-terms for each period (similar results were obtained selecting the top 150 multi-terms for each period). Higher co-occurrences between nodes were grouped forming clusters, distinguishable on the maps as circles of a certain color. To determine these clusters in an automatic way, we followed the same methodology of Testoni et al. [14]. These maps also included as a third depicted variable our general categories’ classification [14]. We plotted the top three general categories associated with each cluster. We inferred that the most frequently connected multi-terms corresponded to those research topics and methodologies that define CONICET’s research agenda in HBMS [14,22].

## Results

A recent article built a proxy of the HBMS prevailing research network defined by the top 200 institutions in terms of frequency of co-publishing in top impact factor journals [14]. CONICET, which is a leading research institution in Latin America (occupying the 183^rd^ and 141^st^ position in the 2019 and 2022 Scimago rankings, respectively), was not part of this HBMS prevailing international research network of organizations. Actually, in our specific analysis, CONICET appeared in the 767^th^ position, far below leading academic institutions and several pharmaceutical corporations, reinforcing our assertion of CONICET as a non-leading research institution (Table 1).

**Table 1.** Ranking of scientific institutions and corporations according to their overall publishing frequency in distinct documents for the chosen corpus. Years: 1999 - 2018. The table displays the top 25 institutions, the top 6 private corporations (red), as well as CONICET’s position (light blue). For each institution or corporation, the number of distinct documents is also shown.

| RANK | INSTITUTION/CORPORATION | NUMBER OF DISTINCT DOCUMENTS |
| --- | --- | --- |
| 1 | HARVARD | 12067 |
| 2 | UNIV CALIF | 11090 |
| 3 | UNIV LONDON | 5598 |
| 4 | UNIV TEXAS | 4874 |
| 5 | UNIV WASHINGTON | 4801 |
| 6 | UNIV STANFORD | 3624 |
| 7 | NIH | 3462 |
| 8 | JOHNS HOPKINS UNIV | 3256 |
| 9 | UNIV OXFORD | 2968 |
| 10 | MAX PLANCK | 2831 |
| 11 | MIT | 2600 |
| 12 | UNIV COLUMBIA | 2529 |
| 13 | UNIV TORONTO | 2474 |
| 14 | UNIV CAMBRIDGE | 2432 |
| 15 | DUKE UNIV | 2419 |
| 16 | UNIV PENN | 2377 |
| 17 | MEM SLOAN KETTERING CANC CTR | 2300 |
| 18 | UNIV YALE | 2194 |
| 19 | UNIV MICHIGAN | 2171 |
| 20 | UNIV CORNELL | 2043 |
| 21 | MAYO CLIN | 1889 |
| 22 | UNIV COLORADO | 1733 |
| 23 | MT SINAI HEALTH SYSTEM | 1718 |
| 24 | UNIV COPENHAGEN | 1705 |
| 25 | UNIV CHICAGO | 1611 |
| 87 | ROCHE | 820 |
| 125 | NOVARTIS | 633 |
| 161 | GLAXOSMITHKLINE | 496 |
| 183 | PFIZER | 435 |
| 189 | AMGEN INC | 420 |
| 198 | MERCK | 402 |
| 767 | CONICET | 74 |

We thus assessed the most prevalent terms found in titles, keywords, and abstracts in the corpus of CONICET HBMS scientific publications between 1999 and 2018. Similar to what was found for the international HBMS agenda [14], multi-terms linked to molecular and cellular biology hegemonize CONICET’s agenda. More than 60 percent of the most frequent multi-terms in different documents were associated with these categories (Table 2). Again, like in the international HBMS agenda (but less prevalent), multi-terms associated with cancer research (approximately 4% of all multi-terms) represent the category with the strongest presence in relation to a specific group of pathologies in both sub-periods (Table 2). However, this prevalence is about three times lower than in the international HBMS agenda. Similarly, research in cardiovascular diseases is also around three times lower in the CONICET vs the international HBMS research agenda.

**Table 2.**
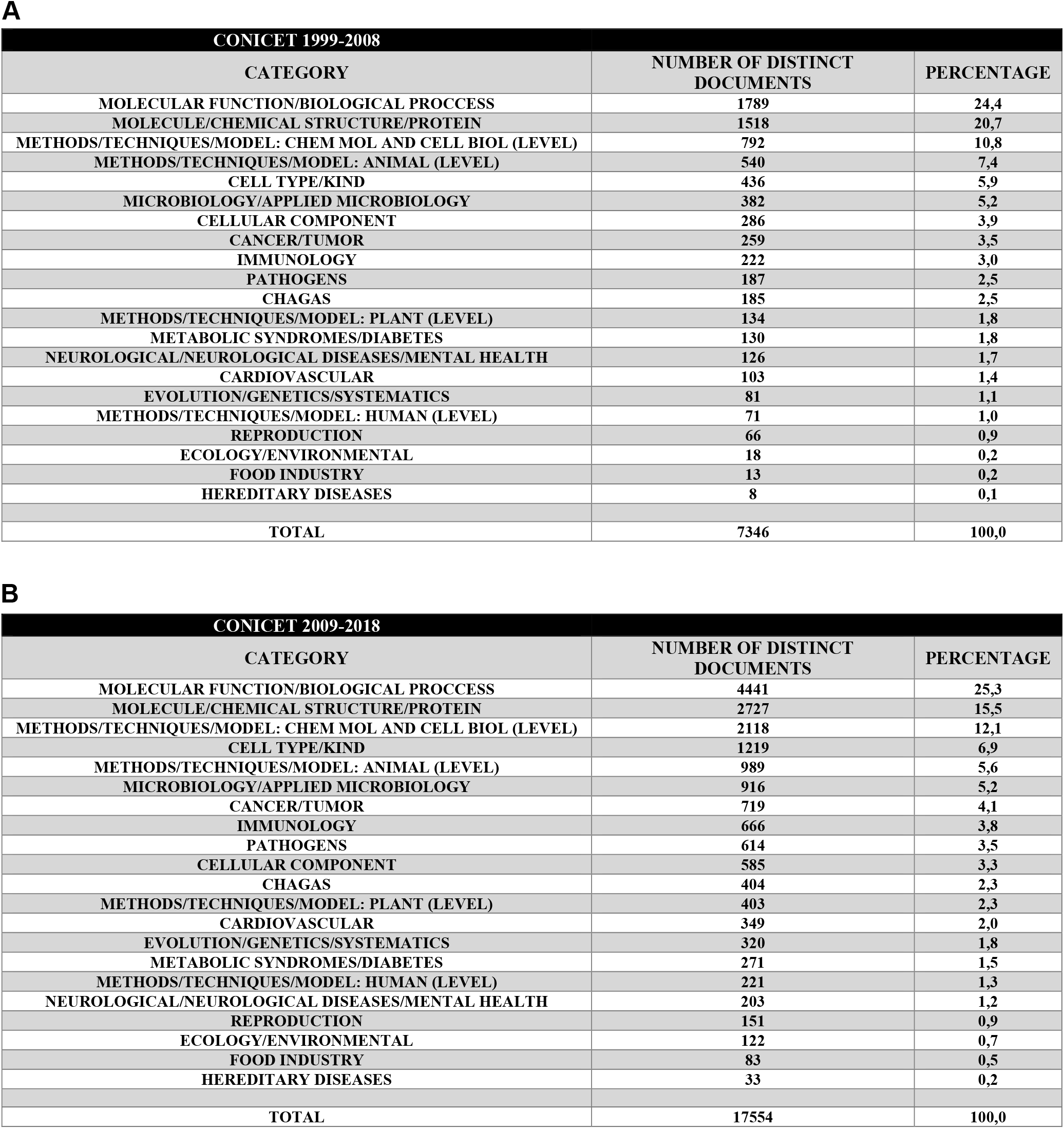
Frequency of multi-terms associated to different categories in the CONICET HBMS agenda. The tables display the accumulated frequency of occurrence of multi-terms corresponding to a particular category. The CONICET HBMS research agenda is divided by period: 1999-2008 (A) and 2009-2018 (B).

While a single cluster integrated by terms related to cancer appears in the first sub-period (Figure 1, light blue circle), a second cluster arises in the second sub-period. One is associated with breast cancer (encompassing terms such as “breast cancer cell” and “estrogen receptor”, among others) and the other one with prostate cancer (Figure 2, light blue and salmon circles, respectively).

**Fig 1.**
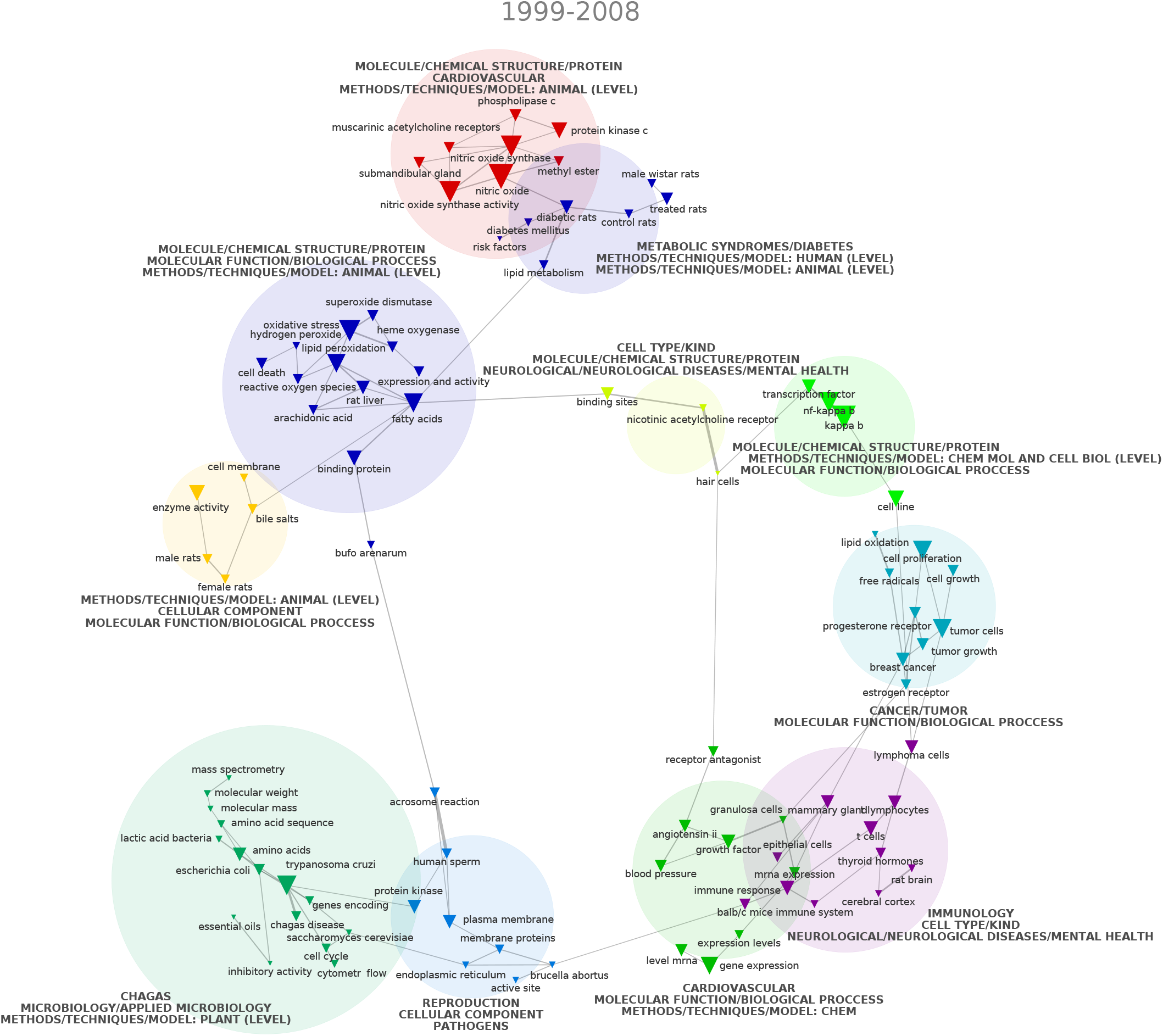
Top 100 CONICET HBMS research multi-terms (1999 to 2008) plotted according to co-occurrence using a chi-squared distribution. Source: authors’ analysis based on WoS data extraction plotted via CorTexT.

**Fig 2.**
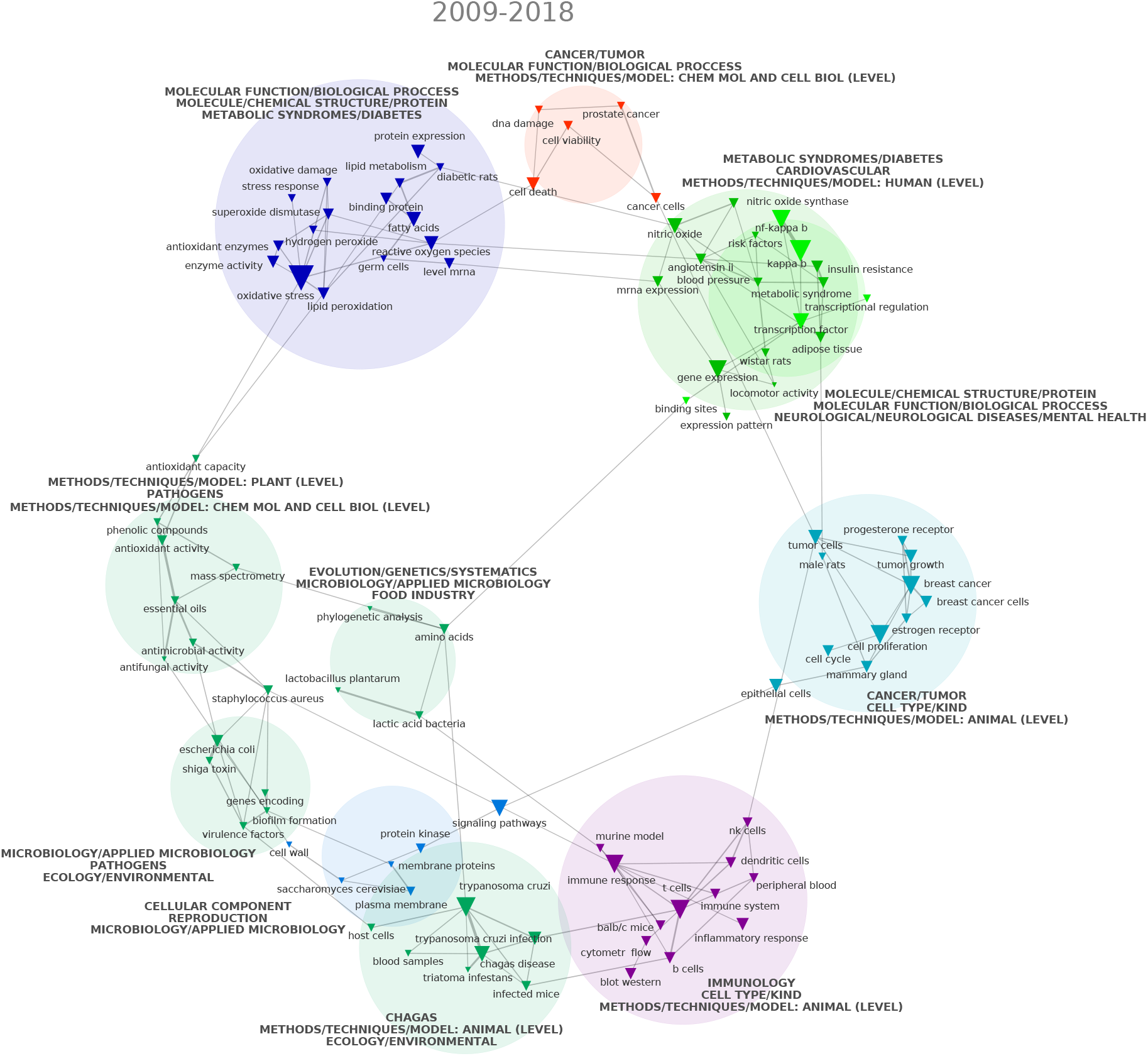
Top 100 CONICET HBMS research multi-terms (2009 to 2018) plotted according to co-occurrence using a chi-squared distribution. Source: authors’ analysis based on WoS data extraction plotted via CorTexT.

Multi-terms related to other specific categories, such as immunology, metabolic disorders, or neurological diseases and mental health show almost the same presence as in the international HBMS agenda (Table 2). Multi-terms connecting health and disease research with ecological, environmental, or social cues were, again as in the international HBMS agenda, marginal. The strong alignment of both agendas for these dominant categories can be observed in Figure 3.

**Fig 3.**
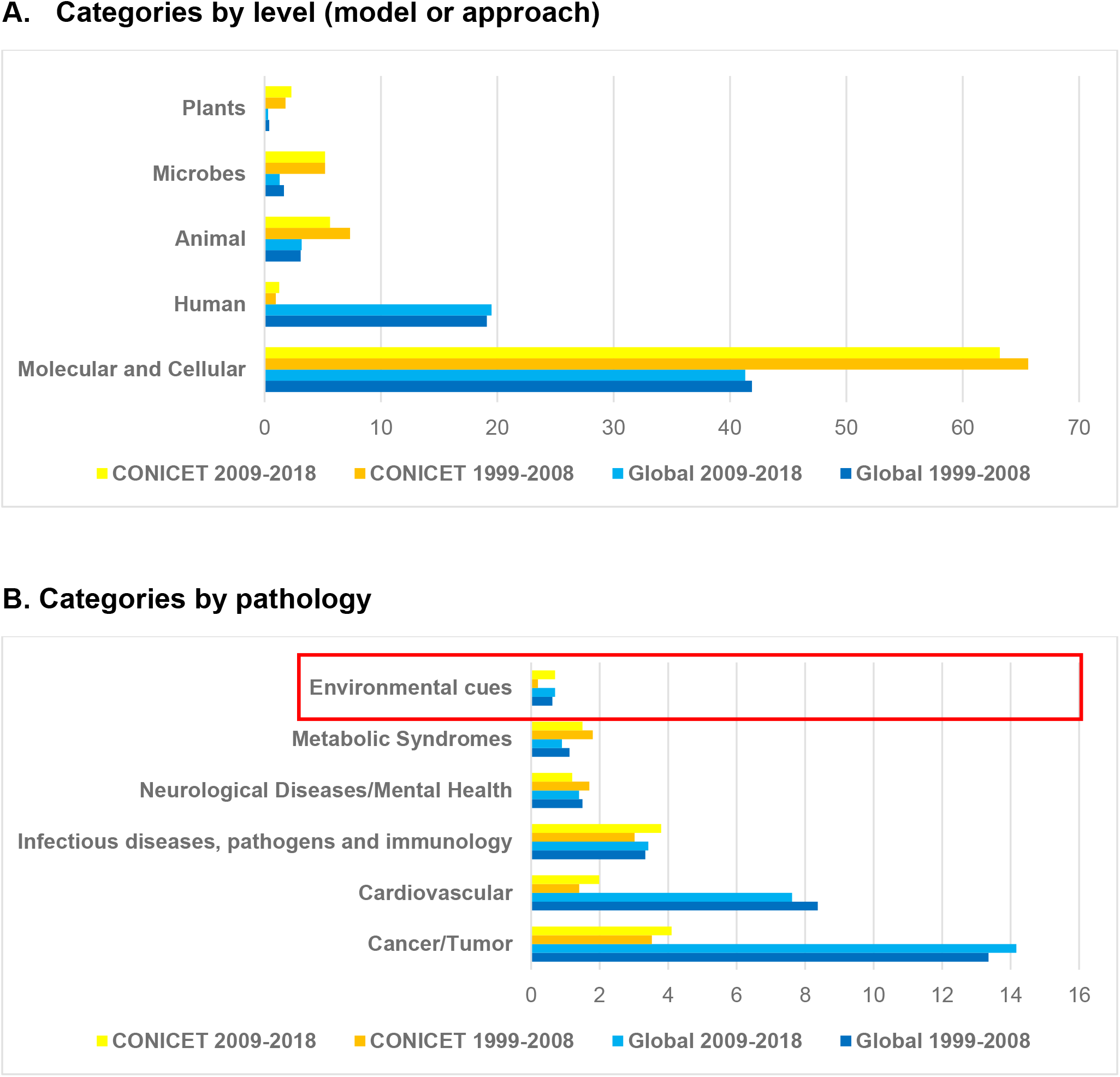
Alignment of the international prevailing and CONICET HBMS agendas. The charts compare the accumulated frequency of occurrence of multi-terms corresponding to different categories in the international and CONICET HBMS agendas. **A**. Categories by organization level (model or approach). **B**. Categories by pathology. The category “Environmental cues” was included in order to compare its relative importance with other categories on the agenda. The agendas are divided by period: 1999-2008 and 2009-2018.

Interestingly, we also found specific differences between the international and CONICET HBMS research agendas. In striking contrast with the former, the presence of multi-terms linked to translational or clinical research (human level) in the latter was negligible. It represented around the one percent of all multi-terms during both sub-periods (about twenty times less than in the international HBMS agenda, Table 2 and Fig. 3). This is consistent with a recent report from the World Health Organization showing that only one percent of global clinical trials occur in Argentina in 2021, as compared to 24% in the US [25].

In turn, CONICET’s HBMS research agenda displayed an increased proportion of multi-terms dealing with categories related to other research levels (models or approaches) -such as molecular and cellular, animals, plants, or microbiology-as compared to the international HBMS research agenda (Fig. 3). Particularly, the enrichment in multi-terms referring to plant research and agrobiotechnology is also reflected in the emergence of a specific cluster in the second period (Figure 2, light green circle). Consistently, this category exhibits a 30 percent increase in multi-terms compared to the first period (Table 2).

Also, as it can be observed in Figures 1 and 2, we found multi-terms associated with pathogens, virulence factors, microbiology, and endemic diseases such as Chagas (“*Trypanosoma cruzi*”, “*Triatoma infestans*”, light green circles), which are marginal in the international agenda. Together, multi-terms related to categories such as microbiology, pathogens, and endemic diseases like Chagas are five to ten times more represented than in the international agenda (Table 2) [14].

## Discussion

Testoni et al. (2021) have recently shown that the prevailing HBMS agenda prioritizes research on pharmacological intervention over research on socio-environmental factors influencing disease onset or progression [14]. Particularly, this research reported that the prevailing HBMS agenda is mostly based on molecular biology methodologies and approaches, with an inclination towards the field of cancer research, neglecting studies on pathogens and biological vectors related to recent epidemics. The authors showed that the actors setting the HBMS prevailing research agenda are simultaneously large private companies and leading academic institutions, whose research is increasingly intertwined.

In this paper, we explored the potential existence of an additional indirect effect exercised by private corporations and leading academic institutions on research institutions from countries that do not participate in HBMS international agenda setting. For this purpose, we analyzed the research agenda of the CONICET as a case study. Our results show that CONICET’s HBMS research agenda, like the prevailing HBMS international research agenda, privileges molecular biology methodologies and approaches and neglects research on the socio-environmental determinants of disease (Fig. 3). These findings apply to both analyzed periods independent of the near three-fold increase in the number of HBMS publications in the second period.

Our findings are consistent with the academic dependency theory [15,18,19] and, particularly, with the emergence of what we call a *scientific dominant discourse* (SDD). In line with the dominant discourse theory proposed by Raiter [24], this concept can be defined as a reference axis that assigns value to the signs in a discursive community (in this case, the HBMS research community), therefore conditioning the circulation of each production (i.e. papers) and the position that certain items occupy in the agenda (HBMS topics and methods). The existence of an SDD implies a disciplinary role during scientific production, thus aligning local research agendas to the international prevailing agenda [26]. Consequently, the SDD regulates the prevalence (or marginalization) of certain research lines. Moreover, if there are economic reasons to privilege certain topics, approaches, methodologies, and sub-fields within a field, it is expected that they will be privileged not only in terms of scientific production but also regarding public and, in particular, private funding. Our results are also consistent with what Lander called “coloniality of knowledge”. The latter is identified as the complex social and epistemological framework that determines the modes of production of science and technology, including which core countries (in particular, which institutions from those countries) concentrate power, domination, and wealth [27].

Nevertheless, in line with Beigel’s qualitative analysis [15,16,28], our results also show that the CONICET’s HBMS research agenda is internally heterogeneous introducing some specific differences in relation to the prevailing HBMS research agenda as depicted by Testoni et al. [14]. Multi-terms associated with categories that include pathogens and endemic diseases are five to ten times more represented than in the international agenda, implying that endemic diseases do receive more attention than in the international HBMS research agenda. These multi-terms integrate clusters with multi-terms related to the field of molecular biology, like “protein kinase” or “molecular weight”. This confirms that molecular biology is the privileged approach in this field, consistent with previous results reflecting that socio-environmental determinants of human health as well as other methodological approaches are relegated [14].

Interestingly, while CONICET’s HBMS agenda shows only a marginal presence of multi-terms linked to clinical or human-level medical research, multi-terms associated with other organization levels or models (such as animal, plant, microbe, or molecular and cellular systems) are over-represented in comparison with the international HBMS research agenda. For instance, multi-terms linked to molecular and cellular biology are even more prevalent in the CONICET HBMS research agenda than in the international one (in which they are already strongly enriched). The same happens with multi-terms related to animal models. This could be explained by a global division of labor where translational medical research, which is sophisticated and expensive, is performed in core countries. In these countries, the cooperation between leading academic and health institutions is robust and dynamic [29]. On the contrary, the institutions from the rest of the world are mostly relegated to suppliers of basic experimental results, which are then exploited by leading academic institutions from core countries as well as by large pharmaceutical corporations.

These differences, far from pointing to local autonomy, are consistent with the theories of coloniality of knowledge and academic dependency in relation to a model of knowledge extractivism. The latter was defined as a process by which science and technology produced by public institutions in the peripheries (or semi-peripheries) is monetized in core countries, generally by companies that monopolize access to knowledge [8,30,31]. A way to measure knowledge extractivism is the identification of blind transfer of knowledge, i.e., the citation of scientific publications in patents where the former’s authors are not among patent co-owners and often are not even aware of the existence of such patents [32,33]. A concrete example of this form of knowledge extractivism concerns COVID-19 related papers. Even though large pharmaceutical companies have benefited heavily from the pandemic, Beall et al. (2021) have recently shown that industry-affiliated articles represented only 2% of worldwide publications on COVID-19 [34].

Another important difference between the international and CONICET’s HBMS prevailing research agendas is the enrichment in multi-terms associated with plant research, agrobiotechnology, and food industry. This is consistent with the leading role of Argentina as a producer of primary goods derived from agrobiotechnology, through the consolidation of its agroindustry during the 1990s [35]. Similarly, the over-representation of multi-terms associated with categories from applied microbiology and food industry in the CONICET HBMS research agenda is consistent with the role of Argentina in the international division of labor as a commodity producer and exporter [36]. Our results are, therefore, coherent with theories arguing that there is a more global model where core countries’ firms capture not only knowledge but also natural resources from the rest of the world, causing significant environmental damage and social conflicts [37–41]. Consistently, the second period shows a relative growth of 3.5 times in the category “Ecology/environmental”, associated with an increase of almost seven times in the number of publications in that category (Table 2 and Fig. 3).

To conclude, CONICET’s HBMS research agenda resembles the prevailing HBMS research agenda. This similarity, nonetheless, presents significant caveats. On the one hand, part of the research privileges certain diseases and favors specific research approaches and methodologies that coincide with those of the HBMS international research agenda depicted by Testoni et al. [14], particularly dominated by the field of molecular biology. On the other hand, our results also show that CONICET’s HBMS research agenda is internally heterogeneous, providing evidence of the co-habitation of heteronomy and a certain degree of autonomy. Yet, the presence of several distinctive research topics (i.e. agrobiotechnology, applied microbiology, and food industry) found in CONICET HMBS research agenda can be explained by the (dependent) place of Argentina in the international division of labor [42,43]. It could be argued that this position influences part of the academic community to dedicate HBMS research efforts on topics that can inform this economic sector’s innovations. In this sense, the research agenda of the CONICET seems to be the result of a combination of elements that reflects a degree of academic and economic dependency [16,44]. In the latter, CONICETs’ distinctive research topics could respond to the action of research groups that build a local or regional position of power that allows them to dispute funding and prestige.

Our contribution maps bibliometric evidence on the invisible network of power relationships that underlies the HBMS international research agenda, indirectly influencing research agendas in the rest of the world. The main contribution of this article is to shed light on the way in which international prevailing research agendas subordinate the public research of a semi-peripheral country. This is consistent with the existence of an SDD which acts as a global axis of reference for publications within the field of HBMS. In the case study analyzed in this work, heteronomy and a certain degree of academic autonomy coexist. However, this apparent autonomy seems to be mostly the expression of another form of dependency, in this case Argentina’s economic dependency, which links the production of knowledge with Argentina’s economic productive structure.

To our knowledge, there are two main limitations to be pointed out. Firstly, network maps do not allow us to assure a causality from the prevailing HBMS research agenda towards CONICET’s agenda. However, their similarity suggests such directionality given the fact that the prevailing HBMS research agenda is set by global leading organizations publishing in the journals with the highest impact factor, while the CONICET has only marginally published in those journals. Secondly, the scope of the WoS may leave behind local publications, with a potentially low international interest, which may be further addressing Argentina’s endemic or specific pathologies.

We understand that from these results a series of questions emerges that it would be interesting to pursue in future investigations. It is relevant to study the potential of research groups rooted outside global leading research institutions to push forward emergent discourses inside their local/national academic community. On the other hand, since the CONICET is a public research institution, it would be interesting to analyze the relation between public policies oriented to set its research agenda and the agenda depicted by our bibliometric maps, considered as proxies of the actual research performed in those years. Finally, it will be interesting to explore the content of other peripheral or semi-peripheral HBMS research agendas, since our work predicts that in addition to the presence of methodologies and approaches related with the SDD, a specific set of topics will be found according to the place that each country occupies in the international division of labor.

## Data Availability

All relevant data are within the manuscript and its Supporting Information files.

## Acknowledgements

We wish to thank Hilary Rose, Steven Rose, Nicolás Rascovan, Pablo Nicolás Fernández Larrosa, Javier Gasulla, Francisco Velázquez Duarte, and Liliana Dain for encouraging discussions that greatly helped to improve our manuscript. We are grateful to Cortext Manager team for their help.

## References

1. Cockburn I, Henderson R. Public–private interaction in pharmaceutical research. Proceedings of the National Academy of Sciences. 1996;93: 12725–12730.

2. Larsen MT. The implications of academic enterprise for public science: An overview of the empirical evidence. Research Policy. 2011;40: 6–19.

3. Lee H, Miozzo M. How does working on university–industry collaborative projects affect science and engineering doctorates’ careers? Evidence from a UK research-based university. The Journal of Technology Transfer. 2015;40: 293–317.

4. Lundh A, Lexchin J, Mintzes B, Schroll JB, Bero L. Industry sponsorship and research outcome. Cochrane Database of Systematic Reviews. 2017.

5. Edwards MG, Murray F, Yu R. Value creation and sharing among universities, biotechnology and pharma. Nature biotechnology. 2003;21: 618–624.

6. Campsall P, Colizza K, Straus S, Stelfox HT. Financial relationships between organizations that produce clinical practice guidelines and the biomedical industry: a cross-sectional study. PLoS medicine. 2016;13: e1002029.

7. Fabbri A, Lai A, Grundy Q, Bero LA. The influence of industry sponsorship on the research agenda: a scoping review. American journal of public health. 2018;108: e9–e16.

8. Rikap C. Asymmetric Power of the Core: Technological Cooperation and Technological Competition in the Transnational Innovation Networks of Big Pharma. Review of International Political Economy. 2019;26: 987–1021. doi:10.1080/09692290.2019.1620309

9. Amorós JE, Poblete C, Mandakovic V. R&D transfer, policy and innovative ambitious entrepreneurship: evidence from Latin American countries. The Journal of Technology Transfer. 2019; 1–20.

10. Albornoz M, Barrere R, Sokil J. Las Universidades Lideran la I+D en América Latina. RICyT. El Estado de la Ciencia 2017. RICyT. RICyT; 2017. pp. 31–44.

11. Arocena R, Göransson B, Sutz J. Knowledge policies and universities in developing countries: Inclusive development and the “developmental university.” Technology in society. 2015;41: 10–20.

12. Arocena R, Sutz J. Weak knowledge demand in the South: learning divides and innovation policies. Science and Public Policy. 2010;37: 571–582.

13. Beigel F. Científicos Periféricos, entre Ariel y Calibán. Saberes Institucionales y Circuitos de Consagración en Argentina. Las publicaciones de los Investigadores del CONICET. Dados. 2017;60: 825–865.

14. Testoni FE, García Carrillo M, Gagnon M-A, Rikap C, Blaustein M. Whose shoulders is health research standing on? Determining the key actors and contents of the prevailing biomedical research agenda. PloS one. 2021;16: e0249661.

15. Beigel F, Gallardo O, Bekerman F. Institutional expansion and scientific development in the periphery: The structural heterogeneity of Argentina’s academic field. Minerva. 2018;56: 305–331.

16. Beigel F. Introduction: Current tensions and trends in the World Scientific System. Sage Publications Sage UK: London, England; 2014.

17. Beigel F. A multi-scale perspective for assessing publishing circuits in non-hegemonic countries. Tapuya: Latin American Science, Technology and Society. 2021;4: 1845923.

18. Alatas SF. Academic dependency and the global division of labour in the social sciences. Current sociology. 2003;51: 599–613.

19. Arvanitis R, Gaillard J. Science indicators for developing countries. International Conference on Science Indicators in Developing Countries, ORSTOM/CNRS, UNESCO, Paris. 1992. pp. 15–19.

20. Kreimer P. Dependientes o integrados?: La ciencia latinoamericana y la nueva división internacional del trabajo. Nómadas. 2006; 199–212.

21. Tancoigne E, Barbier M, Cointet J-P, Richard G. The place of agricultural sciences in the literature on ecosystem services. Ecosystem Services. 2014;10: 35–48.

22. Barbier M, Bompart M, Garandel-Batifol V, Mogoutov A. Textual analysis and scientometric mapping of the dynamic knowledge in and around the IFSA community. Farming Systems Research into the 21st century: The new dynamic. Springer; 2012. pp. 73–94.

23. McCombs M. A Look at Agenda-setting: past, present and future. Journalism Studies. 2005;6: 543–557. doi:10.1080/14616700500250438

24. Raiter A. Discourse Formations and Ideological Reproduction: The Concept of Dominant Discourse. Rethinking Marxism. 1999;11: 87–98.

25. World Health Organization. Number of clinical trials by year, country, WHO region and income group (1999-2019) (Mar 2020). In: Global Observatory on Health Research and Development [Internet]. 2020. Available: https://www.who.int/observatories/global-observatory-on-health-research-and-development/monitoring/number-of-clinical-trials-by-year-country-who-region-and-income-group-mar-2020

26. Raiter A. Capitulo 1. Representaciones sociales. En Al filo de la lengua Medios, publicidad y política Raiter, A y Zullo, J(Comp) Argentina: La Bicicleta Ediciones. 2016; 15–35.

27. Lander E. Eurocentrism and colonialism in Latin American social thought. Nepantla: Views from South. 2000;1: 519–532.

28. Beigel F. A multi-scale perspective for assessing publishing circuits in non-hegemonic countries. Tapuya: Latin American Science, Technology and Society. 2021;4: 1845923.

29. Yao Q, Lyu P-H, Ma F-C, Yao L, Zhang S-J. Global informetric perspective studies on translational medical research. BMC medical informatics and decision making. 2013;13: 1– 15.

30. Rikap C. Becoming an intellectual monopoly by relying on the national innovation system: the State Grid Corporation of China’s experience. Research Policy. 2022;51: 104472.

31. Pagano U. The crisis of intellectual monopoly capitalism. Cambridge Journal of Economics. 2014;38: 1409–1429.

32. Codner DG, Becerra P, Díaz A. Blind technology transfer or technological knowledge leakage: a case study from the south. Journal of technology management & innovation. 2012;7: 184–195.

33. Wang L, Li Z. Knowledge flows from public science to industrial technologies. The Journal of Technology Transfer. 2021;46: 1232–1255.

34. Beall RF, Moradpour J, Hollis A. The private versus public contribution to the biomedical literature during the COVID-19, Ebola, H1N1, and Zika public health emergencies. PloS one. 2021;16: e0258013.

35. Pengue WA. Transgenic crops in Argentina: the ecological and social debt. Bulletin of Science, Technology & Society. 2005;25: 314–322.

36. Leguizamón A. Disappearing nature? Agribusiness, biotechnology and distance in Argentine soybean production. The Journal of Peasant Studies. 2016;43: 313–330.

37. Giarracca N, Teubal M. Argentina: Extractivist dynamics of soy production and open-pit mining. The New Extractivism. 2014; 47–65.

38. Svampa M. Commodities consensus: Neoextractivism and enclosure of the commons in Latin America. South Atlantic Quarterly. 2015;114: 65–82.

39. Svampa M. Neo-extractivism in Latin America: socio-environmental conflicts, the territorial turn, and new political narratives. Cambridge University Press; 2019.

40. Lajmanovich RC, Sandoval M, Peltzer PM. Induction of mortality and malformation in Scinax nasicus tadpoles exposed to glyphosate formulations. Bulletin of environmental contamination and toxicology. 2003;70: 0612–0618.

41. López SL, Aiassa D, Benítez-Leite S, Lajmanovich R, Manas F, Poletta G, et al. Pesticides used in South American GMO-based agriculture: A review of their effects on humans and animal models. Advances in molecular toxicology. 2012;6: 41–75.

42. Ebenau M, Liberatore V. Neodevelopmentalist state capitalism in Brazil and Argentina: chances, limits and contradictionsh. dms–der moderne staat–Zeitschrift für Public Policy, Recht und Management. 2013;6: 17–18.

43. Cooney P. Reprimarization: Implications for the environment and development in Latin America: The cases of Argentina and Brazil. Review of Radical Political Economics. 2016;48: 553–561.

44. Mazzucato M. The entrepreneurial state: Debunking public vs. private sector myths. New York and London: Anthem Press; 2015.

